# Preterm Prelabour Rupture Of Membranes (PPROM) before 23 weeks gestation: A prospective observational study

**DOI:** 10.1101/2023.03.07.23286863

**Authors:** L Goodfellow, A Care, C Curran, D Roberts, M.A. Turner, M Knight, Z Alfirevic

**Affiliations:** Harris-Wellbeing Research Centre, University of Liverpool; Little Heartbeats Patient Support Group; Liverpool Women’s NHS Foundation Trust; National Perinatal Epidemiology Unit, University of Oxford

## Abstract

**Objectives:** Describe infant and maternal outcomes of a national cohort of women with preterm prelabour rupture of membranes (PPROM) under 23 weeks gestation.

**Design:** Prospective national population-based cohort study using the UK Obstetric Surveillance System (UKOSS).

**Setting:** All 194 obstetric units in the UK.

**Participants:** 330 women with singleton and 38 with multiple pregnancies and PPROM between 16^+0^ and 22^+6^ weeks gestation 1/9/19-28/2/21.

**Main outcome measures:** Infant outcomes: livebirth, survival to hospital discharge and severe morbidity, defined as intraventricular haemorrhage grade 3 or 4 and/or supplemental oxygen requirement at 36 weeks postmenstrual age.

Maternal outcomes: surgery for placental removal; sepsis; admission to intensive treatment unit (ITU) and death.

**Methods:** All data including rates of termination of pregnancy for medical reasons (TFMR) were reported. Three rates were calculated for infant outcomes: i) all TFMR excluded; ii) assuming that all TFMR and those with missing data would have died; iii) assuming that all TFMR and those with missing data would be liveborn. Rates are presented as i (ii to iii).

**Results:** For singleton pregnancies the livebirth rate was 44% (30 to 62%), infant survival to discharge was 26% (16 to 54%) and 18% (12 to 49%) of infants survived without severe morbidity. Maternal sepsis rate was 12% for singleton and 26% for twin pregnancies. Surgery for placental removal was 20% and 14%, respectively.

Five women became severely unwell with sepsis, 2 died and a further 3 required ITU care.

**Conclusions:** Although significant numbers of pregnancies with very early PPROM have favourable outcomes, morbidity and mortality rates in this cohort are high for mothers and infants.

These data can be used in counselling families facing PPROM prior to 23 weeks gestation and to underpin research into the complex pathologies, including sepsis, related to this condition. Currently available guidelines should be updated accordingly.

**What is already known on this topic:**

- *PPROM under 23 weeks gestation is a serious pregnancy complication with high rates of morbidity for mothers and infants*
- *Women are often advised to consider termination for medical reasons (TFMR)*
- *Contemporary, population based, pregnancy outcomes are not available, making counselling even more difficult*

**What this study adds:**

- *This study identified significant maternal morbidity; 12% of women developed sepsis and 2 women (0.6%, 95%CI 0.17-2.2%) died*
- *Conversely infant outcomes were relatively favourable; 26% of expectantly managed infants survived to hospital discharge and the potential worst-best case survival range including those that had termination for medical reasons (TFMR) was 16-54%*
- *Understanding of these results are imperative to appropriate counselling and management of women facing this difficult complication*

## Introduction

Preterm prelabour rupture of membranes (PPROM) complicates 30-40% of all preterm births. [1] Serious complications of PPROM include chorioamnionitis leading to maternal and/or neonatal sepsis, placental abruption and stillbirth. UK clinical guidelines exist for management of this condition but only for pregnancies after 24 weeks gestation, i.e. once pregnancy is legally viable.[1] Prior to this gestation, the burden of neonatal morbidity and mortality has been considered so high that termination of pregnancy is generally offered due to extremely low fetal survival and concerns about lifelong neurological disability secondary to extreme prematurity.[2] The incidence of PPROM below 23 weeks gestation is low (∼0.1%), therefore, a typical obstetric unit with 4000 births a year will manage fewer than 5 cases annually.[3] This has led to both a paucity of research and a paucity of clinical experience in expectant management of this condition.

Women and their families report that clinical care in the UK, including offer of termination of pregnancy for medical reasons (TFMR) differs broadly in seemingly similar clinical scenarios.[4] These inconsistencies understandably add to parental distress under already difficult circumstances.

The aim of this study was to provide UK population level data for pregnancies with PPROM between 16^+0^ and 22^+6^ weeks gestation, stratified according to gestation when PPROM occurred. The study was carried out using the UK Obstetric Surveillance System (UKOSS), a research infrastructure that encompasses every consultant-led maternity department in the country.

## Method

### Data collection

The UK Obstetric Surveillance System (UKOSS) is a research platform that collects population-based information about rare pregnancy events from all 194 consultant-led maternity hospitals in the UK. [5]

Nominated reporting clinicians notified UKOSS of all pregnant women who experienced preterm prelabour rupture of membranes between 16^+0^ and 22^+6^ weeks gestation (inclusive). The two exclusion criteria were: pregnancies in which the membranes ruptured before 16^+0^ weeks gestation but were only diagnosed in the 16^+0^ and 22^+6^ week period; and cases where intrauterine death of all fetuses was diagnosed before rupture of membranes. To capture all relevant pregnancies, the minimum latency between PPROM and labour or birth was not specified. Information was requested about all reported pregnancies using a set proforma [6] and regular reminders to return missing data were sent at weeks 6, 10, 14 and 28 after notification, and a final reminder at the end of the data collection period in September 2021. If a woman was still pregnant when the initial data collection form was received then pregnancy outcome data were requested at 2, 6, 10, 14 and 28 weeks after the estimated due date. Referring hospitals that the woman or infant were transferred to were also contacted to request outcome data.

We planned to collect data on pregnancies with PPROM from 1^st^ September 2019 to 31^st^ August 2020, however, when the COVID-19 pandemic was declared in the UK in March 2020 the study was extended for six-months to investigate potential changes in outcomes secondary to the pandemic. Therefore, the study included women with PPROM between 1^st^ September 2019 and 28^th^ February 2021 (inclusive).

### Sample size and statistical analysis

As this was a time limited national observational study no formal power calculation was carried out. The incidence rate was calculated using the denominator of maternities in 2020 from the constituent nations of the UK.[7–9] Statistical analysis was performed in Stata version 15.1 (StataCorp). The study is reported in accordance with the Strengthening the reporting of Observational Studies in Epidemiology (STROBE) statement: guidelines for reporting observational studies.[10]

Demographics are reported for the whole cohort and divided according to whether the pregnancy had expectant management or TFMR. Women who had TFMR after a period of expectant management were included within the TFMR group. Gestational age at PPROM and at birth were calculated according to ultrasound assessment of estimated date of delivery.

Maternal age at estimated date of delivery was calculated assuming a date of birth of 1^st^ June within the given year of birth since only maternal year of birth was collected to maintain anonymity. Body mass index was based on first recorded weight and height in pregnancy. Ethnicity was recorded from medical records, based on the woman’s self-report. Adverse pregnancy history was noted if a woman had previous pregnancy with PPROM between 16^+0^ and 33^+6^ weeks gestation, midtrimester loss between 16^+0^ and 22^+6^ weeks gestation or spontaneous preterm birth (PTB) between 23^+0^ and 36^+6^ weeks.

The impact of the COVID-19 pandemic on pregnancy outcomes was assessed by grouping singleton pregnancies into those with PPROM between 1^st^ September 2019 and 29^th^ February 2020 (‘prior to COVID-19’) and between 1^st^ March 2020 and 28^th^ February 2021 (‘during COVID-19’ pandemic). The rate of reported pregnancies and infant and maternal pregnancy outcomes were compared across the two groups.

The calculated latency between PPROM and birth included all women except those that had a TFMR, thereby including women who had spontaneous births, intrauterine deaths and medically indicated births.

Pregnancy outcomes for infants and mothers are reported separately for singleton and twin pregnancies. Higher order multiples are briefly described. Singleton pregnancy outcomes were divided into four mutually exclusive groups based on when PPROM occurred: 16^+0^-17^+6^ weeks gestation; 18^+0^-19^+6^ weeks gestation; 20^+0^-21^+6^ weeks gestation; and 22^+0^-22^+6^ weeks gestation. The group comprising 22^+0^-22^+6^ weeks gestation was analysed separately because the British Association of Perinatal Medicine (BAPM) produced guidelines in October 2019 suggesting that in some pregnancies, with parental agreement, active resuscitation should be considered at birth from 22^+0^ weeks gestation.[11] Twin pregnancy outcomes are presented according to chorionicity of the pregnancy.

Infant outcomes were: livebirth and survival to hospital discharge with or without severe morbidity defined as intraventricular haemorrhage grade 3 or 4 and/or requirement for supplemental oxygen at 36 weeks postmenstrual age. This outcome was selected in order to allow compatibility with the study by Kibel et al. [13] In addition reporting clinicians were asked to record whether the infant had limb contractures, neonatal seizures, congenital anomalies or severe lung disease during the neonatal course. Severe lung disease was defined as requiring high frequency oscillatory ventilation during the neonatal admission, inhaled nitric oxide during the neonatal admission, or supplemental oxygen at 36 weeks postmenstrual age.

To account for the impact of termination of pregnancy for medical reasons (TFMR) and missing data on the calculated rates of infant outcomes, three rates were calculated: (1) infant outcome in expectantly managed pregnancies with known infant outcome; (2) infant outcome assuming that all pregnancies that had TFMR or an unknown outcome had died; and (3) infant outcome assuming all pregnancies that had TFMR or an unknown outcome had survived.

Details of the type of pregnancy loss that occurred are presented by allocating infants into one of five mutually exclusive groups: birth or intrauterine death before 22^+0^ weeks gestation (often called miscarriage); intrauterine death 22^+0^ or more weeks gestation; neonatal death; TFMR without expectant management; and TFMR after a period of expectant management.

Maternal outcomes were sepsis, surgery for placental removal, intensive treatment unit (ITU) admission and death. All maternal outcomes are reported as defined by local clinicians.

Data are presented as descriptive statistics (mean/median and standard deviation/IQR) and differences between groups were compared using t test for maternal age, Mann-Whitney U test for gestational age at PPROM and Chi squared tests for categorical variables, except for maternal death and ITU admission according to COVID-19 pandemic status which were compared using Fisher’s exact tests. The Wilson score interval was used to generate confidence intervals for proportions where appropriate. A p value <0.05 was considered statistically significant.

### Ethics statement

Ethics committee approval was obtained from the North London REC1 (Ref. Number 10/H0717/20). Cases were reported anonymously by nominated hospital reporting clinicians, and as such consent from patients was neither required nor sought. Further information is available at https://www.npeu.ox.ac.uk/ukoss/completed-surveillance/epprom.

### Patient and public involvement

The patient support and advocacy group, Little Heartbeats, approached the author AC with concerns about inconsistency in counselling and management of cases of PPROM prior to 23 weeks gestation, stimulating this research. CC (the founder of Little Heartbeats) and the patient and public members of the UKOSS Steering Committee were then involved in the design of the study, the conduct of the study and interpretation of the results. CC met regularly with authors LG and AC to review the findings and plan the optimal presentation of the data. The completed analysis was also reviewed by patient and public representation within the UKOSS Steering Committee.

## Results

All 194 UK hospitals participated in UKOSS. One hundred and twenty five (64%) hospitals reported at least one woman with PPROM at 16^+0^-22^+6^ weeks gestation, leading to 551 women reported in total. One hundred and seventy-nine women were removed due to duplication, ineligibility or insufficient information to assess eligibility. Four women were excluded due to false positive diagnosis of PPROM, defined as women in which all follow-up ultrasound scans showed normal liquor volume and birth was at or after 37+0 weeks gestation.

In total 368 women were included in the analysis of whom 38 had multiple pregnancies. There were an estimated 1 011 924 maternities in the UK over 18 months.[7–9] The estimated incidence is therefore 1 case per 2750 maternities (0.04%).

### Demographics

Maternal characteristics were similar when comparing women who had expectant management and those with TFMR (Table 1). Women that had TFMR had an earlier median gestation of PPROM (Table 1).

**Table 1:**
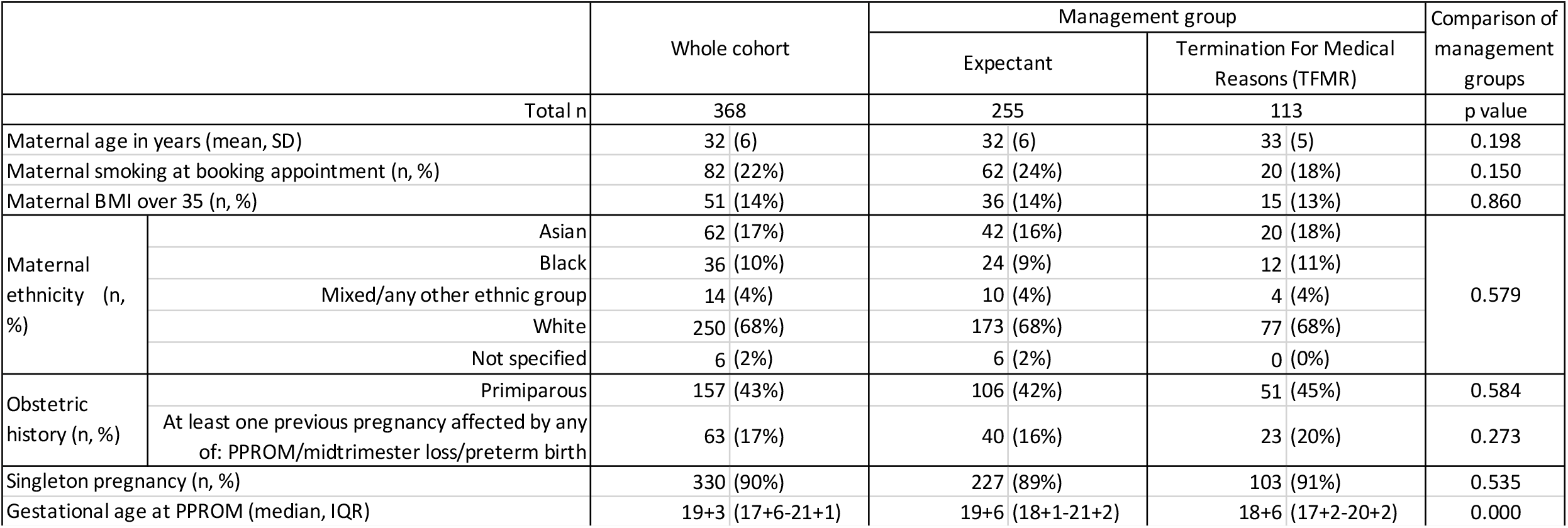
Demographics of cohort, presented both for the whole cohort and according to whether the woman had expectant management or Termination For Medical Reasons (TFMR).

### COVID-19 Pandemic

There were 140 women with PPROM in singleton pregnancies reported to the study in the 6 months prior to the COVID-19 pandemic and 190 in the year during the pandemic. The incidence of reported cases of PPROM was higher prior to the pandemic compared to during the pandemic, with a median of 23 (IQR 19.5-24.5) cases/month and 16.5 (IQR 14-20.25) cases/month respectively (p=0.0004). The number of reported cases per month was lowest for the period July 2020-December 2020 (Figure A1). There were no significant differences detected in infant or maternal pregnancy outcomes according to whether the PPROM occurred prior to or during the pandemic (Tables A1 and A2), therefore the remainder of the analysis was performed using the whole dataset.

### Pregnancy after PPROM

It was possible to calculate the latency between PPROM and birth in 223/227 (98%) of expectantly managed singleton pregnancies. In the immediate period after PPROM the chance of birth was high: 27% (60/223) of births occurred within 72 hours of PPROM and a further 12% (27/223) by 7 days after PPROM (Table 2). Amongst those women that remained pregnant the chance of birth was 21% (29/136) in the second week after PPROM and from the third week onwards the chance of giving birth was approximately 16% of those who remained pregnant per week. This pattern was consistent across the gestational ages of PPROM studied (Figure A2).

**Table 2:** Latency between PPROM and birth in singleton pregnancies with expectant management. Pregnancies that had a termination of pregnancy (n=103) and pregnancies with unknown livebirth status (n=4), are excluded from this table. p value compares latency by gestational age category at PPROM and are calculated by chi squared test.

| Singleton pregnancies with expectant management |  | Whole cohort |  | Gestation at PPROM (weeks) |  |  |  | p value |
| --- | --- | --- | --- | --- | --- | --- | --- | --- |
|  |  |  |  | 16+0-17+6 | 18+0-19+6 | 20+0-21+6 | 22+0-22+6 |  |
| n |  | 223 |  | 43 | 70 | 80 | 30 |  |
| Latency between PPROM and birth | Less than 72 hours | 60 | 27% | 16 (37%) | 18 (26%) | 20 (25%) | 6 (20%) | 0.303 |
|  | 72 hours to <7 days | 27 | 12% | 4 (9%) | 8 (11%) | 9 (11%) | 6 (20%) |  |
|  | 7 days to <28 days | 48 | 22% | 6 (14%) | 12 (17%) | 24 (30%) | 6 (20%) |  |
|  | 28 days or more | 85 | 38% | 17 (40%) | 32 (46%) | 26 (33%) | 10 (33%) |  |
|  | Not specified | 3 | 1% | 0 (0%) | 0 (0%) | 1 (1%) | 2 (7%) |  |

### Infant outcomes

The rate of TFMR declined as gestation of PPROM advanced, from 46% (39/84) of women with singleton pregnancies with PPROM at 16^+0^ -17^+6^ weeks to 19% (7/37) at 22^+0^-22^+6^ weeks (p=0.004, Table 3).

**Table 3:**
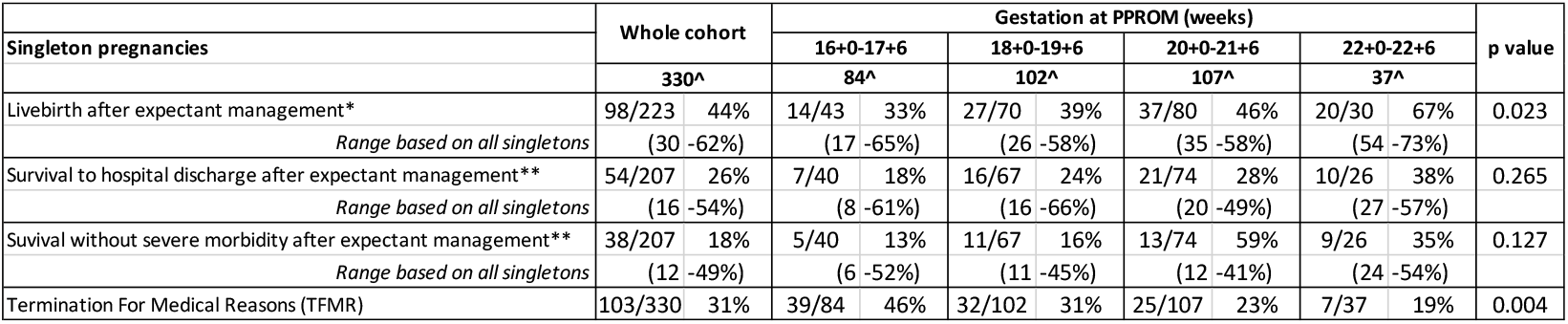
Infant outcomes for singleton pregnancies. p values compare outcomes by gestational age category at PPROM and are calculated by chi squared test. ^Missing data by gestation at PPROM: 16^+0^-17^+6^ weeks livebirth status (2) and discharge status (5); 18^+0^-19^+6^ weeks livebirth status (0) and discharge status (3); 20^+0^-21^+6^ weeks livebirth status (2) and discharge status (8); 22^+0^-22^+6^ weeks livebirth status (0) and discharge status (4) *Excludes pregnancies with termination for medical reasons (TFMR) and those with livebirth status missing ** Excludes pregnancies with TFMR and those with discharge status missing

Amongst the singleton pregnancies that had expectant management and known infant outcome the overall rate of livebirth was 44% (98/223) (Table 3). If one assumes that all pregnancy with TFMR and those with missing data could have been liveborn, the livebirth range would be 62%. In the worst-case scenario with no livebirths in the cohort of pregnancies with TFMR or missing data the livebirth rate estimate is only 30%.

Gestational age at PPROM is an important confounder in expectantly managed pregnancies with the livebirth rate rising from 33% (14/43) in pregnancies with PPROM at 16^+0^ -17^+6^ weeks to 67% (20/30) in pregnancies with PPROM at 22^+0^-22^+6^ weeks (p=0.023) (Table 3).

The overall rate of infant survival to discharge with expectant management was 26% (54/207) when all data needed to assess this outcome were available. The range-based assumption related to TFMR and missing data was 54% (177/330) for the best-case scenario and 16% (54/330) for the worst-case scenario (all TFMR and pregnancies with missing data having an adverse outcome). There was a trend towards improved survival with advancing gestational age at PPROM, but this did not reach our threshold for statistical significance (p=0.265) (Table 3).

The rate of survival to discharge amongst liveborn infants was 55% (54/98). A further 20% (20/98) of liveborn infants had missing data about their discharge status.

The median gestation at birth of liveborn infants was 28^+3^ weeks, interquartile range (IQR) 25^+3^-30^+2^ and the median gestation at birth of infants with known survival to discharge was 29^+4^ weeks, IQR 27^+1^-34^+3^ weeks. The median length of hospital stay after birth for surviving infants was 59 days, IQR 17-100 days.

Amongst the 54 singleton liveborn neonates with known survival to hospital discharge, 16 (30%) met our criteria for severe morbidity with grade 3 or 4 IVH and/or requirement for supplemental oxygen therapy at 36 weeks postmenstrual age (Table 3). The rate of survival without severe morbidity amongst all expectantly managed singleton pregnancies with known outcomes was 18% (38/207). The best-case scenario if infants of all pregnancies with TFMR and missing data had favourable outcomes is a survival to discharge without severe morbidity of 49% (161/330) and the worst-case scenario if all infants of pregnancies with TFMR and missing data died is a survival to discharge without severe morbidity rate of 12% (38/330) (Table 3).

The infant morbidity included grade 3 or 4 IVH in 11% (6/54) of survivors and severe lung disease after birth was reported in 52% (28/54) of surviving infants. Two neonates had limb contractures, affecting one and two limbs respectively and no surviving infants had neonatal seizures. There was no difference in morbidity according to gestational age at PPROM, however the small number of surviving infants with earlier gestations of PPROM limited statistical power. The rate of survival without severe morbidity appeared to be influenced by gestation at birth; the rate for infants born under 28^+0^ weeks gestation was 53% and the rate for those infants born at or after 34^+0^ weeks gestation was 87% (Table A3). The unadjusted relative rate ratio of survival without severe morbidity was 1.23 (95% CI 1.04-1.47, p=0.027) per additional week of gestation at birth.

### Singleton pregnancy losses

Amongst the 310 singleton pregnancies with known discharge status, a total of 256 (83%) resulted in pregnancy loss. Details of the type of pregnancy loss are given in Table 4.

**Table 4:**
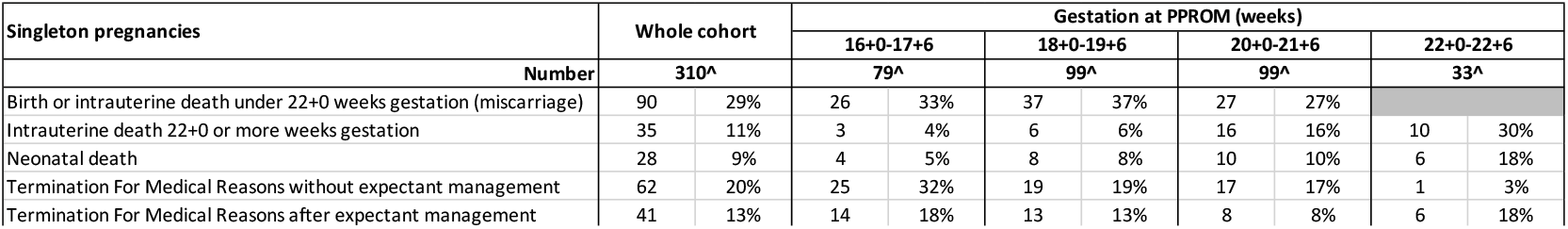
Types of pregnancy loss in singleton pregnancies ^Pregnancies with missing discharge status are excluded: (5) after PPROM at 16^+0-^17^+6^ weeks; (3) after PPROM at 18^+0^-19^+6^ weeks; (8) after PPROM at 20^+0^-21^+6^ weeks; (4) after PPROM at 22^+0^-22^+6^ weeks

The rate of TFMR without expectant management reduced from 32% (25/79) when PPROM occurred at 16^+0^-17^+6^ weeks to 3% (1/33) with PPROM at 22^+0^-22^+6^ weeks gestation. The rate of TFMR after expectant management was relatively similar at 8-18% across gestations studied (Table 4). When PPROM occurred prior to 22^+0^ weeks the rate of birth or intrauterine death under 22^+0^ weeks (often called miscarriage) was relatively consistent at 27-37%. There were more intrauterine deaths at 22^+0^ or more weeks gestation and neonatal deaths when PPROM occurred between 22^+0^ and 22^+6^ weeks gestation (30% and 18% respectively), compared to 9% and 8% respectively with PPROM prior to 22^+0^ weeks gestation (p<0.001 and p=0.05 respectively). This was largely because 39% of pregnancies ended within a week of PPROM (Table 2) which meant that babies born after PPROM at 22^+0^ weeks gestation had a higher chance of birth with extreme prematurity.

### Maternal outcomes

Amongst the 268 women with singleton pregnancies who chose initial expectant management 12% (33/268) developed maternal sepsis (Table 5). Three of these women became severely unwell; two died and a third was admitted to ITU and survived. These three women also required surgical removal of the placenta. Two of the three women deteriorated within 5 days of diagnosis of PPROM, and the third over a month after PPROM.

**Table 5:** Maternal complications of singleton pregnancies. Data for whole cohort presented as n, % of total and (95% confidence interval). Data by gestational age at PPROM presented as n and % of total. p values compare outcomes by gestational age category at PPROM and are calculated by chi squared test. *Whether, or not, the mother developed sepsis was only asked for women who initially opted for expectant management. Therefore, these are expressed as a proportion excluding those with immediate TFMR. Women that had TFMR after expectant management are included, therefore n=268. Surgery for placental removal is expressed as a proportion of all women.

| Maternal outcomes of singleton pregnancies | Whole cohort |  |  | Gestation at PPRM (weeks) |  |  |  |  |  |  |  | p value |
| --- | --- | --- | --- | --- | --- | --- | --- | --- | --- | --- | --- | --- |
|  |  |  |  | 16+0-17+6 |  | 18+0-19+6 |  | 20+0-21+6 |  | 22+0-22+6 |  |  |
|  | 330 |  |  | 84 |  | 102 |  | 107 |  | 37 |  |  |
| Maternal sepsis* | 33 | 12% | (8-16%) | 7 | 12% | 10 | 12% | 10 | 11% | 6 | 17% | 0.801 |
| Surgery for placental removal | 65 | 20% | (15-24%) | 19 | 23% | 23 | 23% | 17 | 16% | 6 | 16% | 0.570 |

The rate of maternal sepsis was not assessed amongst women that had TFMR without expectant management (n=62), however there were no deaths or ITU admissions among these women. The rate of maternal death with singleton pregnancies is therefore 2/330, 0.61%, 95%CI 0.17-2.2%.

Twenty two percent (51/227) of women with singleton pregnancies and expectant management required surgery for placental removal. The rate of surgery for placental tissue removal was 14% (14/103) amongst women with singleton pregnancies who had a TFMR. This difference did not reach statistical significance (p=0.06).

### Multiple pregnancies

In our cohort there were 38 women with multiple pregnancies (10%, 38/368), which is an over-representation as fewer than 2% of births nationally are from multiple pregnancies.[9] Twenty three were dichorionic-diamniotic twins (DCDA), 10 were monochronic-diamniotic twins (MCDA), one was trichorionic triplets. Chorionicity was not determined in four twin pregnancies.

In six out of 30 twin pregnancies with expectant management, both infants survived to hospital discharge (20%). In a further five twin pregnancies (17%) one baby survived to discharge from hospital. (Table 6). If one assumes that all infants with TFMR may have been liveborn, and those with missing information about hospital discharge did survive, then the survival to hospital discharge rate could be as high as 27% for both infants and as high as 49% for a single twin infant at discharge. In the worst-case scenario with no livebirths in the cohort of pregnancies with TFMR and all those with missing discharge status having died then the survival to hospital discharge rate would be 16% for both infants with an additional 14% of pregnancies having a liveborn single infant surviving to discharge. The majority of twin survivors were from DCDA pregnancies (Table 6). Six twin pregnancies (6/37, 16%) had death of a single infant (either intrauterine death or spontaneous birth under 22^+0^ weeks gestation) and TFMR of a second twin.

**Table 6:**
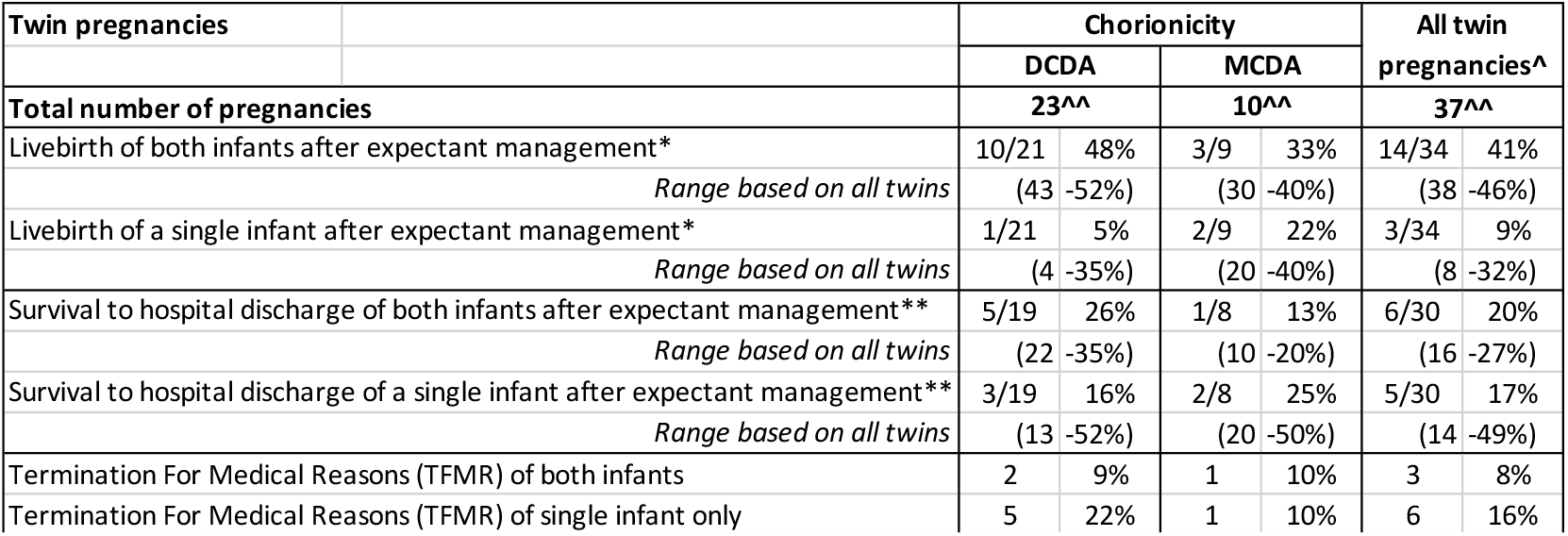
Infant outcomes for twin pregnancies. ^ There were four twin pregnancies with undocumented chorionicity, these are included within the ‘All twin pregnancies’ data only ^^Missing data by chorionicity: DCDA discharge status for both infants (1); DCDA discharge status for a single infant (1); MCDA discharge status for a single infant (1); Unknown chorionicity discharge status for a single infant (1). Livebirth status was available for all twin infants. *Excludes pregnancies with termination for medical reasons (TFMR) of both infants ** Excludes pregnancies with TFMR of both infants and discharge status missing for either or both infants

Amongst the 10 women with MCDA pregnancies there was only one pregnancy with survival of both infants, and a further two pregnancies with survival of a single infant. Importantly, six out of 10 MCDA pregnancies had either laser coagulation or amnioreduction for twin-to-twin transfusion syndrome prior to PPROM.

Thirty women with twin pregnancies (16 DCDA, 8 MCDA and 4 with chorionicity not determined) had expectant management but three (1 DCDA, 1 MCDA and 1 with chorionicity not determined) did not have complete information on infant morbidity. In the cohort with complete data, 17 out of 54 infants (31%) survived until discharge from hospital (6 sibling pairs and 5 single twins). Four of the 17 surviving infants (24%), all from DCDA pregnancies, had severe morbidity when discharged.

The rate of survival to hospital discharge without severe morbidity after expectant management of twin pregnancies was therefore 13/54 (24%) of infants (4 sibling pairs and 5 single twins).

### Maternal morbidity in twin pregnancies

Overall, maternal morbidity in twin pregnancies was similar to that in singleton pregnancies (Tables 5 and 7). However, maternal sepsis was somewhat higher in women with twin pregnancies that had expectant management or TFMR after expectant management (26% compared to 12% for singletons; p=0.03). Two women with multiple pregnancies required intensive care treatment.

**Table 7:** Maternal outcomes of twin pregnancies. ^ There were four twin pregnancies with undocumented chorionicity, these are included within the ‘All twin pregnancies’ data only *Whether, or not, the mother developed sepsis was only asked for women that initially opted for expectant management. Therefore, these are expressed as a proportion excluding those with immediate TFMR (n=35 for all women with twin pregnancies). Women who had TFMR after expectant management are included. The remaining outcomes are expressed as a proportion of all women.

| Maternal outcomes of twin pregnancies | Chorionicity |  |  |  | All twin pregnancies^ |  |
| --- | --- | --- | --- | --- | --- | --- |
|  | DCDA |  | MCDA |  |  |  |
|  | 23 |  | 10 |  | 37 |  |
| Maternal sepsis* | 7 | (32%) | 1 | (11%) | 9 | (26%) |
| Surgery for placental removal | 4 | (17%) | 1 | (10%) | 5 | (14%) |

## Discussion

### Statement of principal findings

The results of this national, population-based study of PPROM prior to 23 weeks gestation starkly illustrate the diverse infant and maternal outcomes possible with this condition.

Whilst 26% (54/207) of women with expectantly managed singleton pregnancies had an infant that survived to hospital discharge, only 18% of women (38/207) had an infant that survived without severe morbidity. Maternal sepsis developed in 12% (33/268) of women with singleton pregnancies and two women died. There is additional complexity because of the uncertainty concerning infant outcomes inherent in 31% of women having TFMR.

### Comparison with previous studies

The two maternal deaths equate to a rate of 606 per 100 000 maternities with PPROM 16^+0^-22^+6^ weeks gestation, 95% CI 166-2183 per 100 000. This strikingly higher than the baseline UK maternal mortality rate of 11 per 100 000 maternities[12] and requires contextualising within the wider scientific literature.

In the past decade over 700 women have been included in observational cohorts of PPROM at a similar gestations with no maternal deaths reported.[3,13–18] This concurs with no maternal deaths reported in the most recent review on the topic,[19] and only a single maternal death reported in the largest review, published in 2009, citing a publication from 1988.[2,20] Therefore the maternal deaths are likely to be surprising to many clinicians. The key difference between the current work and previous studies is that our cohort was population based as we surveyed all 194 consultant-led maternity units in the UK in contrast to previously published studies from five or fewer (often specialised) centres. In accordance with the population-based approach second trimester PPROM associated with maternal sepsis and death has featured in three of the UK’s maternal mortality reports in the past decade. [25–27] Population level data has also identified seven women who died after PPROM between 14^+0^ and 24^+6^ weeks gestation from 2001 to 2015 in France, giving an estimated chance of death of 45 per 100 000 maternities with this complication.[21] These women had similar causes of death to the current cohort; six were attributed to sepsis and one to haemorrhage secondary to placenta accreta spectrum. Therefore, whilst the absolute risk of maternal death with very early PPROM is likely to be within the lower range of our confidence intervals the population-based literature suggests that these are not isolated incidents, and may have previously been under-recognised because the literature has been based on data from a small number of centres.

The rate of maternal infectious morbidity in previous studies of expectantly managed singleton pregnancies is similar to our 12%.[3,13,22] In this cohort maternal sepsis was higher in twin pregnancies, also in concordance with previous work.[23] Surgery for placental removal is a less well recognised complication of birth after PPROM but occurred in 20% of our singleton cohort, including the three women who became severely unwell.

This rate concurs with a cohort of midtrimester PPROM cases in Ireland.[3] The combination of requiring surgery for placental removal and sepsis should alert clinicians to the possibility of maternal deterioration.

The infant survival to hospital discharge rate of 26% for expectantly managed singleton pregnancies, and the range of possible survival of 16-54% when pregnancies with TFMR and unknown outcomes are accounted for are broadly in keeping with observational studies over the past decade, that reported rates of 17-40% with similar gestations of PPROM.[13,15,24–28] Amongst the infants that survived to hospital discharge, 70% avoided severe morbidity by our definition, this is also in keeping with recently published observational studies.[13,22,26,27]

The severe neonatal morbidity definition was chosen for consistency with Kibel *et al* (2016) who related neonatal outcomes at hospital discharge following pregnancies complicated by PPROM at 20-24 weeks gestation to outcomes at a corrected age of 18 – 21 months. Of 24 neonates with grade 3 or 4 IVH and/or requirement for oxygen at 36 weeks postmenstrual age and/or grade 3 or more retinopathy of prematurity, 8 (30%) had moderate-severe morbidity when they were 18-21 months old. Among 27 babies who were born following pregnancies complicated by PPROM at 20-24 weeks gestation but who did not have any IVH, ROP or oxygen requirement outcomes at 36 weeks, none had moderate-severe morbidity when they were 18-21 months old.[22]. Our results are likely to be indicative of long-term moderate-severe morbidity in a similar proportion of children.

Within this study women that had expectant management after PPROM at 16^+0^-17^+6^ weeks gestation and gave birth after 23 weeks gestation had comparable livebirth and infant morbidity rates to those who experienced PPROM at 22^+0^-22^+6^ weeks gestation. This could be due to lack of statistical power because only seven infants survived to discharge after PPROM at 16^+0^-17^+6^ weeks gestation. It is also possible that when PPROM occurred earlier in pregnancy more pregnancies with less favourable characteristics had a TFMR, negating any negative effect of gestation of PPROM on infant morbidity. However a study from The Netherlands, with a rate of termination of only 2%, also found no difference in infant morbidity after at PPROM at ≥13-<20 weeks (n=21) compared to ≥20-<24 weeks (n=41).[26] We therefore suggest that the impact of gestation when PPROM occurs on infant morbidity needs further evaluation.

The median gestational age at birth of surviving infants was 29^+4^ weeks gestation, and the median length of hospital stay after birth for surviving infants was 59 days, IQR 17-100 days. Whilst this is shorter than contemporary studies from Australia [15] (median 76 days, IQR 44-111 days) and Japan [29] (mean 155 days, standard deviation (SD) 53 days), it is still a significant amount of time and likely to have a substantial impact upon the whole family in the medium term. We suggest this information should be included in patient counselling.

As gestational age at birth advanced the chance of livebirth and infant survival to hospital discharge improved, as expected.[2] However, even among those born after 34 weeks gestation two infants (2/15, 13%) had severe morbidity, illustrating the complexities of these cases and the need for ongoing multidisciplinary team care, including neonatologists, even at relatively advanced gestations.

Infants of DCDA multiple pregnancies with PPROM appeared to have comparable pregnancy outcomes to singletons. This is in keeping with previous literature.[30] MCDA pregnancies had lower infant survival, but 60% of these pregnancies also had pathologies of monochorionicity such as twin to twin transfusion syndrome and selective growth restriction. Therefore, the pathologies unique to monochorionic pregnancies are likely to be the key contributor to the mortality in these instances.

### Strengths and Limitations

The UKOSS infrastructure has enabled the largest population-based study of PPROM prior to 23 weeks’ gestation. There is, however, unavoidable uncertainty concerning infant outcomes due to 31% of our population opting for TFMR and inability to follow up all infants, particularly those that moved hospital as part of their care.

The 0.04% incidence of PPROM between 16^+0^-22^+6^ weeks may be an under estimation due to the possibility of under-reporting of cases, particularly during the covid-19 pandemic. However, we have no evidence of biased reporting which might influence the generalisability of the results. A recent French analysis of hospital episode statistics for the whole nation found a prevalence of 0.2% of PPROM 14^+0^-24^+6^ weeks, and the authors considered this figure to be likely an under-estimation, noting that it is impossible with coded data to fully validate the diagnosis.[21] The number of cases reported was lowest between July 2020 and December 2020, possibly because of staffing pressures, or potentially due to a lower rate of PPROM secondary to public health control measures such as lockdowns.

Our survey did not include questions about whether mothers who opted straight for termination of pregnancy after PPROM developed sepsis, and to simplify case reporting and capture the maximum number of cases the inclusion criteria were kept brief and questions regarding fetal anomalies, whether identified at the time of PPROM or not, were not included. The survey reporters were not asked whether women that opted for a TFMR, or had a baby loss, also had a fetus with a life-threatening anomaly. As the presence of such a fetal anomaly may have been a possibility in some instances, their presence may have influenced the decision to terminate the pregnancy. A limited range of neonatal outcomes, and no outcomes after hospital discharge, were collected because of resource constraints.

### Implications for clinical practice

This study illustrates that women with PPROM prior to 23 weeks gestation have some of the highest risks of infant and maternal morbidity that a clinician will face amongst the women they care for. An individual obstetrician is likely to only see one or two women with extremely preterm PROM a year, thereby being unable to build a wealth of experience relevant to the condition. In accordance with other recent studies we also highlight that women opting for TFMR do not avoid all severe morbidity.[23,31]

In current UK practice pregnant women prior to 20 weeks gestation are often cared for on gynaecology rather than obstetric wards. The infant mortality that occurred was largely attributable to birth prior to viability and extreme prematurity because 39% of women gave birth in the week after PPROM, and a further 21% gave birth in the following week. This pattern was consistent across the gestational ages of PPROM studied, and is consistent with previous work. [13,26] We suggest that pregnancies with PPROM under 23 weeks gestation have a high likelihood of needing support from infant bereavement teams, neonatal teams and/or maternal critical care. Obstetricians with an interest in this condition, alongside dedicated midwives, are likely to be best placed to co-ordinate these elements of care in the immediate aftermath of PPROM and for the remainder of the pregnancy. Neonatal teams can note that about half of babies who were admitted to neonatal units following PPROM between 16^+0^-22^+6^ weeks survived. A majority of survivors did not have the morbidities we captured. Among survivors, the likelihood of the morbidities we captured was lower with each week of completed pregnancy.

We suggest that teams with expertise in management of very early PPROM, along with patient representatives, work together to develop guidelines for care.

### Implications for research

This study provides baseline data on infant and maternal outcomes of pregnancies with PPROM under 23 weeks gestation upon which studies aiming to improve outcomes can be planned. Interventions may be novel treatments aimed at treating the pathology, or care bundles, including training, to optimise care and delivery timing using interventions already available.

Materials for women and families facing very early PPROM will need to consider the inherent uncertainty within the data secondary to 31% of women with singleton pregnancies opting for TFMR. The optimal way to communicate such wide uncertainties within the data to a wider audience, and support families with such complex pregnancies, is yet to be determined.

The current study is only able to comment on severe morbidity in infants at discharge from hospital, and previous work suggests that 70% of these infants will not have significant morbidity when aged 18-21 months.[22] The rate of long term disability in offspring is the most desired information for prospective parents. Future research needs to incorporate ways of capturing this information.

There is likely to be substantial unmeasured psychological morbidity related to this condition. The optimal way of managing very early PPROM to support psychological wellbeing of families requires further consideration.

## Conclusions

Women with PPROM prior to 23 weeks gestation can have favourable maternal and pregnancy outcomes. However, a significant proportion of these pregnancies are complicated by maternal morbidity and infant mortality and morbidity. All clinicians who care for these families need to be conscious of the risk of maternal sepsis and death. The data presented will be helpful in counselling families facing PPROM prior to 23 weeks gestation and should be incorporated into updates of clinical guidelines.

## Data Availability

Data cannot be shared publicly because of confidentiality issues and potentially identifiable sensitive data as identified within the Research Ethics Committee application/approval. Requests to access the data can be made by contacting the National Perinatal Epidemiology Unit data access committee via.

## Acknowledgements

We would like to acknowledge the assistance of UK Obstetric Surveillance System reporting clinicians, the Little Heartbeats volunteers and the UK Obstetric Surveillance System Steering Committee without whose support this research would not have been possible. We would also like to thank Mrs Tracey Ricketts for administrative assistance.

## Transparency statement

LG (the manuscript’s guarantor) affirms that the manuscript is an honest, accurate, and transparent account of the study being reported; that no important aspects of the study have been omitted; and that any discrepancies from the study as originally planned have been explained.

