## Supplementary material for "Preterm Prelabour Rupture Of Membranes (PPROM) before 23 weeks gestation: A prospective observational study": A1

### Appendix 1 for UKOSS PPROM

#### COVID-19 Pandemic

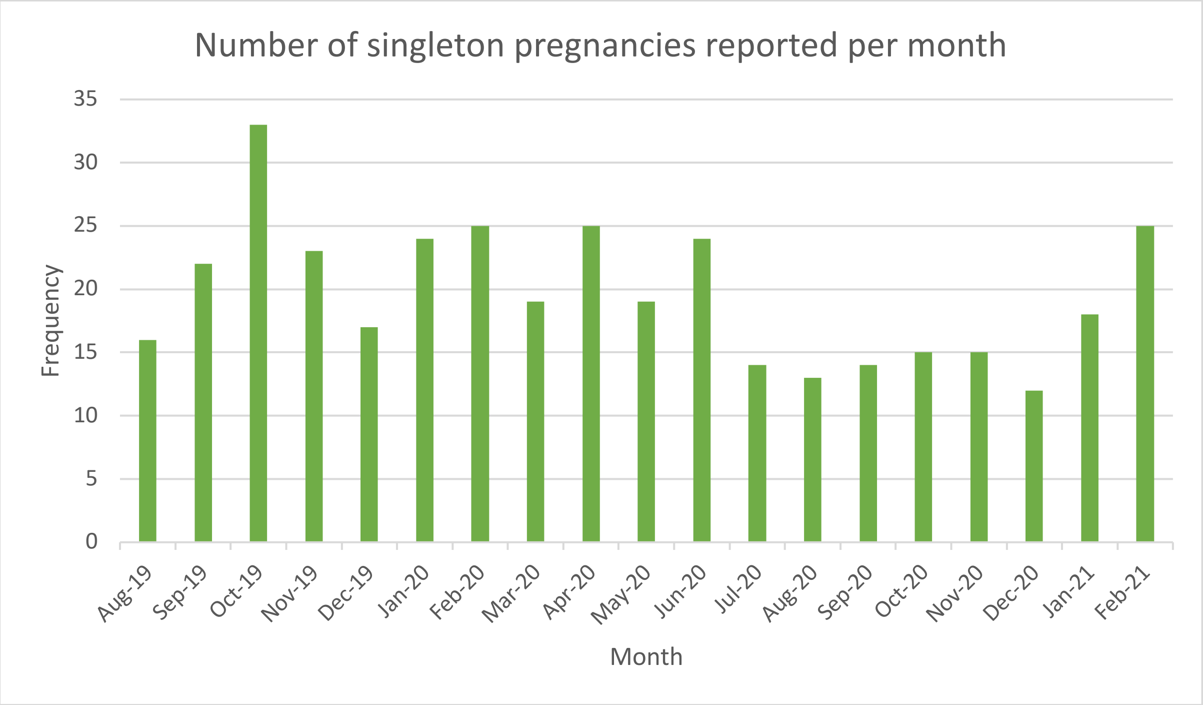


*Figure A1: Number of singleton pregnancies reported to the study by month*

| Infant outcomes of all singleton pregnancies | Prior to COVID-19 pandemic | | During COVID-19 pandemic | | P value |
|---|---|---|---|---|---|
| **Number** | **140** | | **190** | | |
| Termination of pregnancy | 45 | (32%) | 58 | (31%) | 0.434 |
| Pregnancy loss, stillbirth, or livebirth with neonatal death (n, %) | 65 | (46%) | 88 | (46%) | |
| Livebirth with survival to hospital discharge (n, %) | 23 | (16%) | 31 | (16%) | |
| Livebirth with unknown discharge status (n, %) | 4 | (3%) | 12 | (6%) | |
| Unknown pregnancy outcome (n, %) | 3 | (2%) | 1 | (1%) | |

*Table A1: Infant outcomes of singleton pregnancies according to whether preterm premature rupture of membranes occurred prior to or during the COVID-19 pandemic (which commenced in March 2020).*

| Maternal outcomes of all singleton pregnancies | Prior to COVID-19 | | During COVID-19 | | P value |
|---|---|---|---|---|---|
| **Number** | **140** | | **190** | | |
| Sepsis | 13 | (9%) | 20 | (11%) | 0.548 |
| Surgery for placental removal | 26 | (19%) | 39 | (21%) | 0.900 |
| ITU admission | 0 | (0%) | 2 | (1%) | 0.510 |
| Death | 0 | (0%) | 2 | (1%) | 0.510 |

*Table A2: Maternal outcomes of singleton pregnancies according to whether preterm premature rupture of membranes occurred prior to or during the COVID-19 pandemic (which commenced in March 2020).*

#### Pregnancy after PPROM

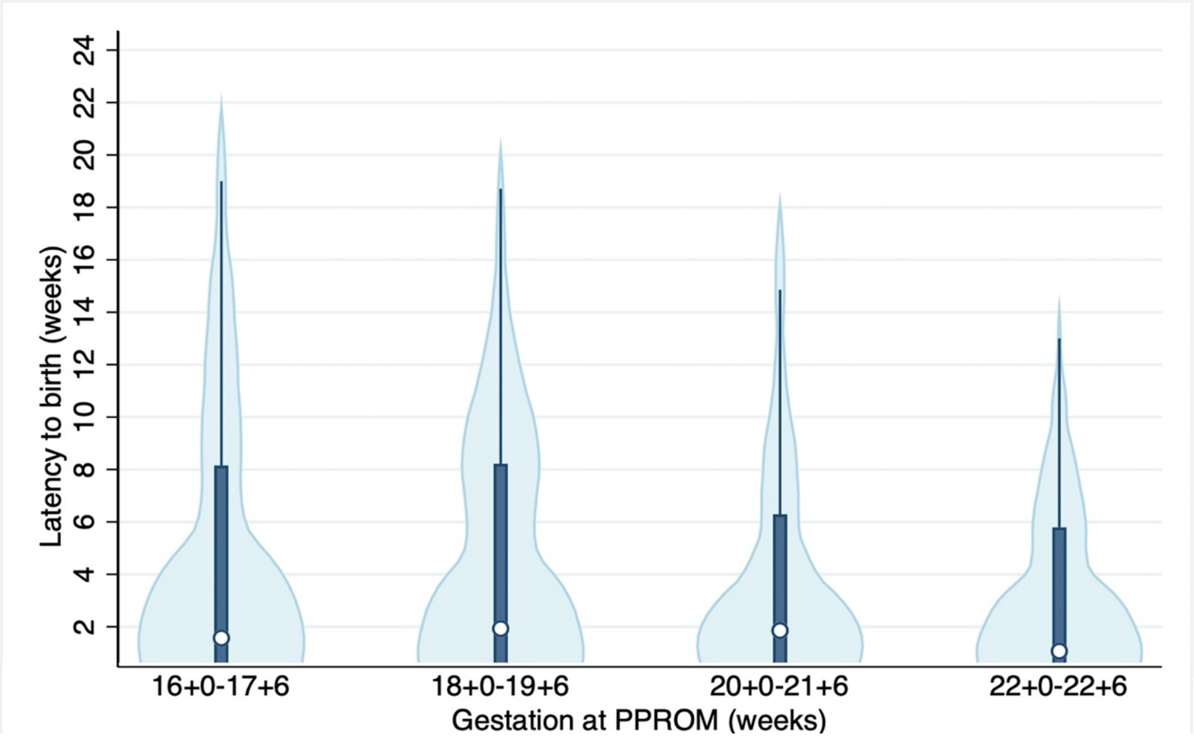


*Figure A2: Violin plot illustrating the latency between PPROM and birth for women with conservative management and singleton pregnancies (n=223). The white dot shows the median value (1 week 6 days for the whole group) the dark bar shows the IQR (3 days to 7 weeks 1 day for the whole group). The thin blue line shows 1.5x the interquartile range. The light blue area represents the kernel density estimation to show the distribution shape of the data. The plot has been truncated at 0 weeks.*

#### Severe morbidity by gestational age at birth

| Percentatge of surviving infants that did not have severe morbidity n=54 | **Gestation at birth** | | | | | | | | | | | |
|---|---|---|---|---|---|---|---|---|---|---|---|---|
| | **Under 24 weeks** | | **24+0-27+6 weeks** | | **28+0-31+6 weeks** | | **32+0-33+6 weeks** | | **34+0-36+6 weeks** | | **37+0 or over** | |
| Survival without severe morbidity | 1/4 | 25% | 7/11 | 64% | 15/22 | 68% | 2/2 | 100% | 10/12 | 83% | 3/3 | 100% |
| | (0 | -67%) | (35 | -92%) | (49 | -88%) | | | (62 | -100%) | | |

*Table A3: Percentage of surviving infants that did not have severe morbidity by gestational age at birth in singleton pregnancies. Presented as n/N, % and 95% confidence interval*
